# Low risk of SARS-CoV-2 transmission via fomite, even in cold-chain

**DOI:** 10.1101/2021.08.23.21262477

**Authors:** Julia S. Sobolik, Elizabeth T. Sajewski, Lee-Ann Jaykus, D. Kane Cooper, Ben A. Lopman, Alicia NM. Kraay, P. Barry Ryan, Jodie L. Guest, Amy Webb-Girard, Juan S. Leon

## Abstract

**Background:** Countries continue to debate the need for decontamination of cold-chain food packaging to reduce possible SARS-CoV-2 fomite transmission among workers. While laboratory-based studies demonstrate persistence of SARS-CoV-2 on surfaces, the likelihood of fomite-mediated transmission under real-life conditions is uncertain.

**Methods:** Using a quantitative risk assessment model, we simulated in a frozen food packaging facility 1) SARS-CoV-2 fomite-mediated infection risks following worker exposure to contaminated plastic packaging; and 2) reductions in these risks attributed to masking, handwashing, and vaccination.

**Findings:** In a representative facility with no specific interventions, SARS-CoV-2 infection risk to a susceptible worker from contact with contaminated packaging was 2·8 × 10^−3^ per 1h-period (95%CI: 6·9 × 10^−6^, 2·4 × 10^−2^). Implementation of standard infection control measures, handwashing and masks (9·4 × 10^−6^ risk per 1h-period, 95%CI: 2·3 × 10^−8^, 8·1 × 10^−5^), substantially reduced risk (99·7%). Vaccination of the susceptible worker (two doses Pfizer/Moderna, vaccine effectiveness: 86-99%) combined with handwashing and masking reduced risk to less than 1·0 × 10^−6^. Simulating increased infectiousness/transmissibility of new variants (2-, 10-fold viral shedding) among a fully vaccinated workforce, handwashing and masks continued to mitigate risk (2·0 × 10^−6^ -1·1 × 10^−5^ risk per 1h-period). Decontamination of packaging in addition to these interventions reduced infection risks to below the 1·0 × 10^−6^ risk threshold.

**Interpretation:** Fomite-mediated SARS-CoV-2 infection risks were very low under cold-chain conditions. Handwashing and masking provide significant protection to workers, especially when paired with vaccination.

**Funding:** U.S. Department of Agriculture

## 1. Introduction

According to the WHO^1^ and U.S. Centers for Disease Control and Prevention,^2^ fomite-mediated transmission of SARS CoV-2 is rare.^3-7^ This is especially in the context of overall SARS-CoV-2 transmission and relative to the predominant aerosol and droplet transmission modes.^8-10^ Fomite-mediated transmission can occur when an uninfected individual transfers infectious particles from a contaminated surface to their facial mucosa, typically via their hands.^11^ These surfaces can become contaminated from an infected individual by: 1) shedding onto hands which then touch a surface; or 2) expelled respiratory aerosols and droplets (coughs, speaks, sneezes etc.,)^12,13^ which then fall to a surface.^14^ However, definitive epidemiological evidence of fomite transmission is lacking. Only a few case reports implicate fomites as a possible SAR-CoV-2 source ^15-17^ of which, asymptomatic aerosol transmission could not be eliminated as an alternative transmission mode (aerosol, droplet, fomite-mediated).

Despite these sparse epidemiological data, a recent report of the isolation of infectious SARS-CoV-2 from a single environmental sample of imported frozen cod packaging in Qingdao, China ^18^ has raised alarm for fomites as viral vectors for SARS-CoV-2 transmission, particularly in the context of seeding SARS-CoV-2 into areas that have controlled transmission.^19^ Exacerbating this concern, laboratory studies suggest prolonged infectivity (days to perhaps weeks) of SARS-CoV-2 ^20^ on a variety of surfaces ^11,21-23^ and low temperatures and humidity (common in cold-chain conditions) are associated with enhanced virus stability (months or longer).^24^ In addition, SARS-CoV-2 viral RNA has been detected on surfaces in playgrounds and water fountains,^25^ high-touch community fomites,^26^ on surfaces in households with asymptomatic patients,^27^ and on surfaces in close proximity to infected individuals in healthcare settings.^28-36^ However, the relationship between detectable viral RNA and infectious virus is tenuous.^37^ Of an identified 63 published studies testing for SARS-CoV-2 RNA from environmental surface samples, only 13 attempted to isolate infectious virus. Of these, viable SARS-CoV-2 virus was identified in only four instances: frozen cod packaging,^18^ a nightstand within a household of a COVID-19 confirmed case,^38^ an isolation room of patients undergoing mechanical ventilation,^39^ and on a windowsill of a patient’s quarantine unit.^40^ In the context of a cold-chain occupational setting, evidence is lacking on the frequency of SARS-CoV-2 contamination on food packaging and the association between its detection by RT-qPCR and infectious virus.

The relative importance of fomite-mediated transmission as investigated by modeling studies is also inconclusive, especially across diverse settings. Using a quantitative microbial risk assessment (QMRA) framework, several studies involving contact with community-based fomites estimated SARS-CoV-2 infection risks to be low (on the order of 1 in 10,000).^26,41,42^ In occupational settings, relative risks associated with fomite transmission of SARS-CoV-2 were higher (range: 2 × 10^−2^ – 3·2 × 10^−1^ infection risks) in child daycare centers,^43^ hospitals,^44,45^ and a food manufacturing facility.^46^ Higher still was the mathematical modeling of the Diamond Princess cruise ship outbreak ^47^ which suggests a 30% attribution of COVID-19 cases to fomites.

In an effort to prevent and control potential SARS-CoV-2 outbreaks associated with imported food products, China has implemented procedures to test all imported cold-chain (temperature-controlled transport and storage) products and their packaging, including disinfection and wet wiping ^48^ of plastic packaging.^19^ However, there is no definitive evidence of SARS-CoV-2 fomite transmission from contact with contaminated food or food packaging,^5^ suggesting that these decontamination measures may be extreme.^4-7,49^ Given this, there is a need to inform the frozen and cold-chain food industries on the possible risk of SARS-CoV-2 contamination of, and transmission from, food packaging, under cold-chain conditions, and the impact of infection control measures. In this study, a stochastic QMRA model is used to quantify the risks and relative risk reductions attributed to fomite-mediated SARS-CoV-2 transmission and infection control measures among workers in a representative frozen food packaging facility under cold-chain conditions. We also investigate the need for additional plastic packaging decontamination.

## 2. Materials and Methods

### 2.1 Model overview

Quantitative microbial risk assessment (QMRA) is a mathematical modeling framework used to evaluate health risks associated with direct and indirect transmission pathways and the efficacy of infection control strategies. For this work, we applied the validated QMRA model of Sobolik, *et al*.,^46^ to simulate contamination of plastic packaging (individual cartons and plastic-wrapped palletized cartons) with respiratory particles via the coughing of two infected workers. SARS-CoV-2 exposure doses and infection risks resulting exclusively from fomite transmission were simulated for a susceptible worker in a receiving warehouse.

### 2.2 Model structure

The overall model structure initiated with two infected workers in a representative frozen food manufacturing facility (Figure 1). The first worker was located within close proximity (≤3 feet) of a conveyor belt which transported between 144 and 216 individual plastic cartons (dimensions: [0·38m × 0·28m × 0·15m] or [0·38m × 0·30m × 0·23m]) per hour. The second infected worker transferred these individual plastic cartons onto a wooden pallet (36-54 cartons/pallet), either manually or by automation, and then wrapped the pallet in plastic wrap (four pallets processed/hour). Contamination events occurred through expelled aerosol (<50 µm) and droplet (50-750 μm) respiratory particles generated from cough events by the two infected workers. Plastic wrapped, palletized cartons were then transported under cold-chain storage to a receiving warehouse where a single susceptible worker was exposed to the virus exclusively by direct contact with contaminated plastic wrap and/or surface-contaminated individual plastic cartons during manual unpacking of the pallets.

**Figure 1.**
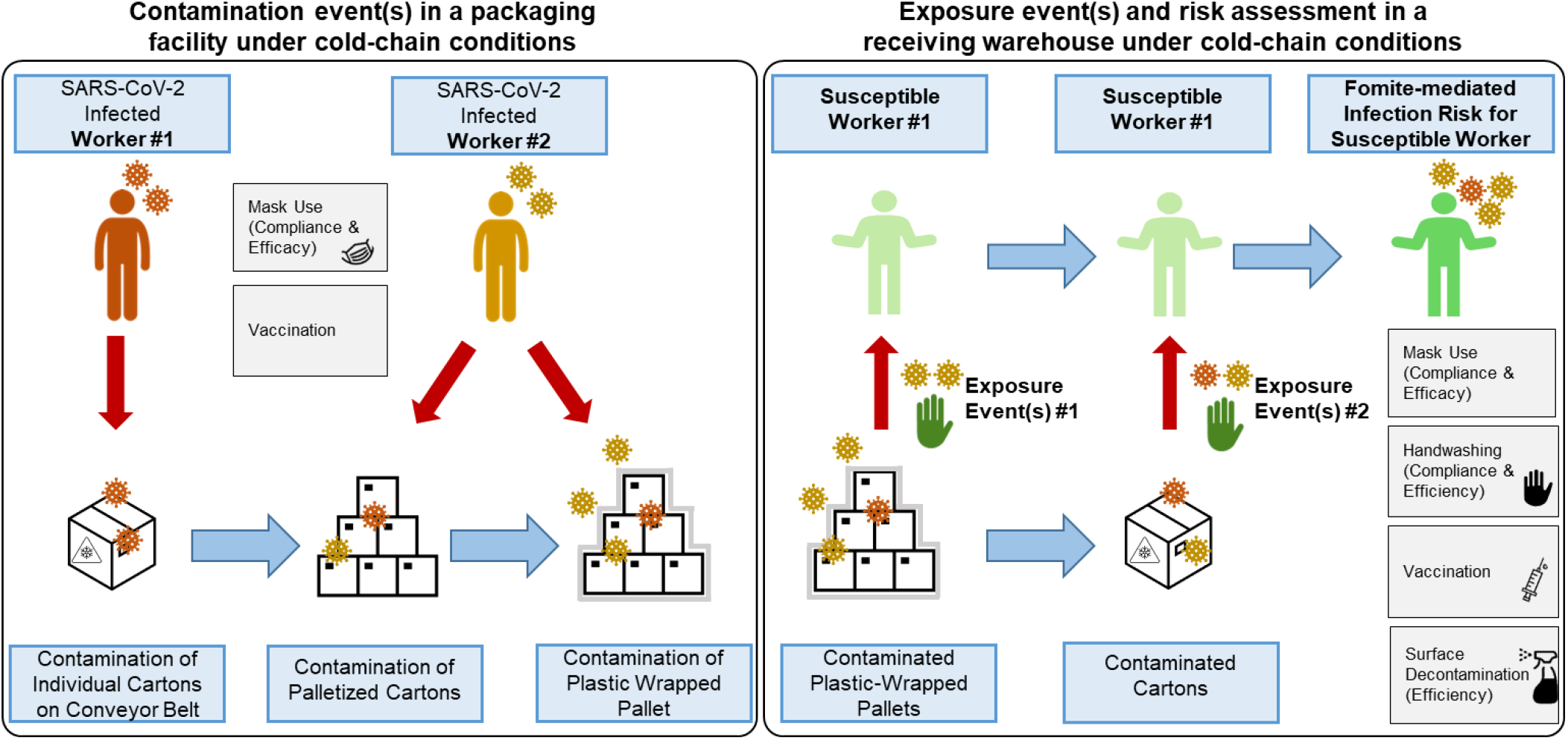
Conceptual framework for fomite-mediated SARS-CoV-2 transmission involving exposure of a susceptible worker to individual plastic cartons, palletized cartons, and plastic wrap in a receiving warehouse under cold-chain conditions. This schematic depicts a representative frozen food packaging facility, initiating with two infected workers (left panel). Up to 10 contamination events per infected worker (0 to 10 coughs) can occur at three stages in the packaging pipeline: 1) contamination of the top-face of individual plastic cartons (144-216 individual cartons processed per hour) via respiratory droplet and aerosol fallout from the first infected worker while cartons are transported along a conveyor belt (orange in schematic); 2) contamination of cartons via respiratory particle spray (droplets and aerosols) as cartons are placed (manually or via automation) on a pallet by the second infected worker (yellow in schematic); and 3) contamination of the plastic-wrapped palletized cartons by respiratory particle spray (droplet and aerosol) from the second infected worker (yellow in schematic). Four pallets, each containing approximately 36-54 individual plastic cartons, are processed per hour. Because of current Good Manufacturing Practices (cGMP), the model did not account for indirect transfer of virus from the infected workers’ hands to the plastic fomites along the packaging pipeline. Under cold-chain conditions assuming no viral decay, plastic wrapped pallets were transported to a receiving warehouse for unloading by a susceptible worker. Infection risks resulting exclusively from fomite transmission were simulated as contacts between the susceptible worker’s fingers and palms (of both hands) and the fomite surface (accounting for the surface area of the hand relative to the fomite surface); virus transfer from fomite to hands; and virus transfer from fingertips to facial mucous membranes (accounting for the surface area of the fingers relative to the combined surface area of the eyes, nose, and mouth). Grey boxes indicate infection control measures implemented for the infected (mask use, vaccination) and susceptible (handwashing, mask use, vaccination) workers. In the scenarios with additional plastic surface decontamination, this was simulated prior to the susceptible worker contacting the fomites.

The two model outcomes included: 1) SARS-CoV-2 infection risks from fomite-mediated exposures to plastic packaging (individual plastic cartons and plastic wrapped pallets) following a 1-hour period; and 2) relative reduction in SARS-CoV-2 infection risk attributed to infection control interventions (masking, handwashing, vaccination, and package surface decontamination). The model was developed in R (version 4.0.3; R Development Core Team; Vienna, Austria) using the mc2d package for Monte Carlo simulations.^50,51^ For each simulation, 10,000 iterations were run. Please refer to *SI Appendix* for additional details on model assumptions, vetting, and sensitivity analyses.

### 2.3 Data sources

Model parameters were derived from the peer-reviewed literature (see *SI Appendix*, Table S1) and included: (i) viral shedding through cough events; (ii) fomite-mediated transmission parameters; (iii) dose-response parameters for SARS-CoV-2 infection risk; and (iv) risk mitigation interventions.

### 2.4 Fomite-mediated transmission modeling

SARS-CoV-2 contamination of the individual plastic cartons was calculated using the combined aerosol and droplet particle fallout, Fall_t,a_ (infectious virus) and Fall_t_ (infectious virus/m^3^), expelled from coughs by the first infected worker as described in Sobolik *et al*.^46^ Contamination of the palletized cartons and the plastic wrapped-pallets was calculated using the combined aerosol and droplet particle spray, C_t,aerosol_ (PFU/m^3^) and C_t,droplet_ (PFU/m^3^), expelled from coughs by the second infected worker as previously described ^46^ with the resulting viral contamination on the fomite surfaces, Fomite_cartons_ and Fomite_plasticwrap_ (PFU/m^2^):

Viral concentration on individual cartons (PFU/m^2^):

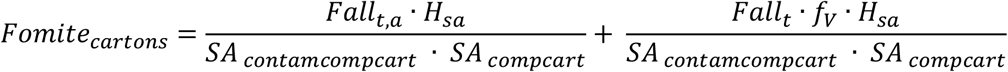

Viral concentration on plastic wrap pallets (PFU/m^2^):

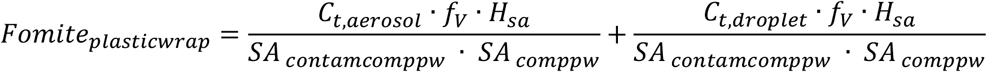

where f_V_ was the air volume of the facility (m^3^), H_sa_ was the surface area of the susceptible worker’s hand that touched the fomite surface (m^2^), SA_contamcompcart_ was the cross-sectional area of the composite contaminated individual cartons (m^2^), SA_compcart_ was the cross-sectional area of the composite individual cartons (m^2^), SA_contamcomppw_ was the cross-sectional area of the contaminated plastic wrap (m^2^), and SA_comppw_ was the cross-sectional area of the composite total plastic wrap (m^2^). Homogenous mixing of aerosol particles was assumed when calculating the aerosol contamination of the individual cartons and plastic-wrapped pallets. The ability of droplets (50-750 μm) to contaminate fomites as fallout or spray was determined by their size, particle transport properties (i.e. ballistic gravitational trajectories), and distance traveled. The proportion of droplets that reached the plastic cartons (droplet fallout) or plastic wrap (droplet spray) within 0 to 3 feet distancing was derived from previous modeling work.^52^

To calculate the concentration of SARS-CoV-2 transferred to a hand, C_hand,carton_ (PFU/h), following contact with the individual cartons, Fomite_cartons_ (PFU/ m^2^), we applied an approach previously use for influenza A virus exposure.^53^

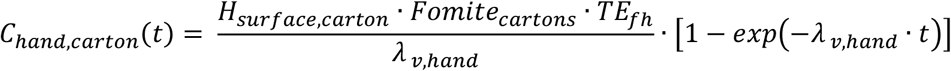

Where H_surface,carton_ was the frequency of contacts between the hand and the cartons per minute (contacts/min), Fomite_cartons_ was the viral concentration on the cartons (PFU/m^2^), at time t, TE_fh_ was the proportion of virus transferred from fomite to hand, and λ_v,hand_ was the viral decay of SARS-CoV-2 on the hand. Similarly, we calculated the concentration of SARS-CoV-2 transferred to a hand, C_hand,pw_ (PFU/h), following contact with the plastic wrap:

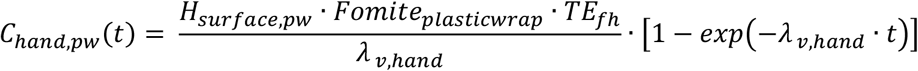

Where H_surface,pw_ was the frequency of contacts between the hand and plastic wrap per minute (contacts/min), Fomite_plasticwrap_ was the viral concentration on the plastic wrap fomite (PFU/m^2^), at time t, TE_fh_ was the proportion of virus transferred from fomite to hand, and λ_v,hand_ was the viral decay of SARS-CoV-2 on the hand.

### 2.5 Risk assessment

The fomite-mediated dose to the susceptible worker following contact while unloading the palletized cartons was calculated from the viral contamination on the hand (C_hand,carton,_ C_hand,pw_) at time t, the frequency of hand-to-face contacts (H_face_), the surface area of the hands (H_sa_), the surface area ratio of fingers (F_sa_) to face (Face_sa_), the fraction of pathogens transferred from hand-to-face (TE_hm_), and the exposure duration (t):

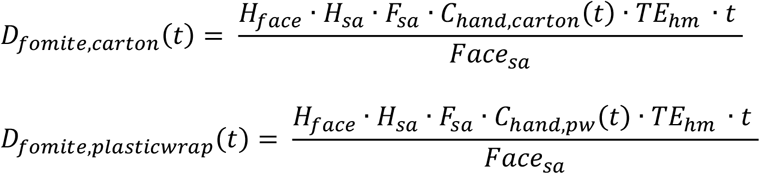

The total viral dose, *D*_*fomite,total*_, (PFU) at time t:

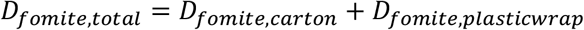

In the absence of a SARS-CoV-2 dose-response model, the probability of SARS-CoV-2 infection to the susceptible worker was calculated by applying an exponential dose-response model (k_risk_) (PFU^-1^) based on pooled data from intranasal administration of SARS-CoV and murine hepatitis virus on infection in mice,^54,55^ and consistent with other SARS-CoV-2 QMRA models.^26,41,42,44^

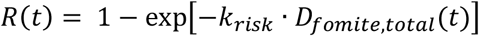

### 2.6 Evaluation of risk mitigation interventions

Standard infection control measures were selected based on current OSHA, CDC, and FDA-recommended guidance and industry practices for worker safety^56^ and COVID-19 prevention.^57^ These interventions included concordant masking (surgical) and hourly handwashing (2 log_10_ virus removal).^58^ Vaccination (two doses of Pfizer/Moderna) was implemented with and without the standard infection control measures to reduce 1) susceptibility to infection of the worker in the receiving warehouse (susceptible worker vaccinated only); and 2) overall transmissibility when all workers are vaccinated (rare breakthrough infection among vaccinated workers). Please refer to *SI Appendix* for additional details on the vaccination scenarios. Further, we simulated the added effect of surface decontamination (3 log_10_ virus removal)^48,59^ applied directly to plastic packaging (individual cartons, plastic wrap) as described elsewhere,^19,48^ combined with the standard infection control measures (handwashing, masking). While there are no reference risk thresholds for respiratory pathogens, all fomite-mediated risks were compared to the reference ranges of 1·0 × 10^−4^ and 1·0 × 10^−6^ risks, which are WHO and U.S. EPA thresholds for water quality for representative infectious diseases.^60,61^

## 3. Results

### 3.1 Fomite-mediated SARS-CoV-2 infection risks associated with exposures to individual plastic cartons and plastic wrap during the unpacking of products under cold-chain conditions, in a receiving warehouse

In the scenario of no vaccination/prior infection, the risk of fomite-mediated transmission without standard infection control measures was 2·8 × 10^−3^ per 1h-period (95%CI: 6·9 × 10^−6^, 2·4 × 10^−2^) (Figure 2A). Implementing standard infection control measures substantially reduced risk by 66·7% for masking (9·4 × 10^−4^ risk per 1h-period, 95%CI: 2·3 × 10^−6^, 8·1 × 10^−3^), by 99.0% for handwashing (2·8 × 10^−5^ risk per 1h-period, 95%CI: 6·9 × 10^−8^, 2·4 × 10^−4^), or by 99·7% for handwashing and masks (9·4 × 10^−6^ risk per 1h-period, 95%CI: 2·3 × 10^−8^, 8·1 × 10^−5^), relative to no infection control measures. The addition of plastic surface decontamination to these infection control measures reduced infection risks by 100% to below the 1 × 10^−6^ risk threshold (9·39 × 10^−9^ risk per 1h-period, 95%CI: 2·3 × 10^−11^, 8·1 × 10^−8^), relative to no infection control measures.

**Figure 2.**
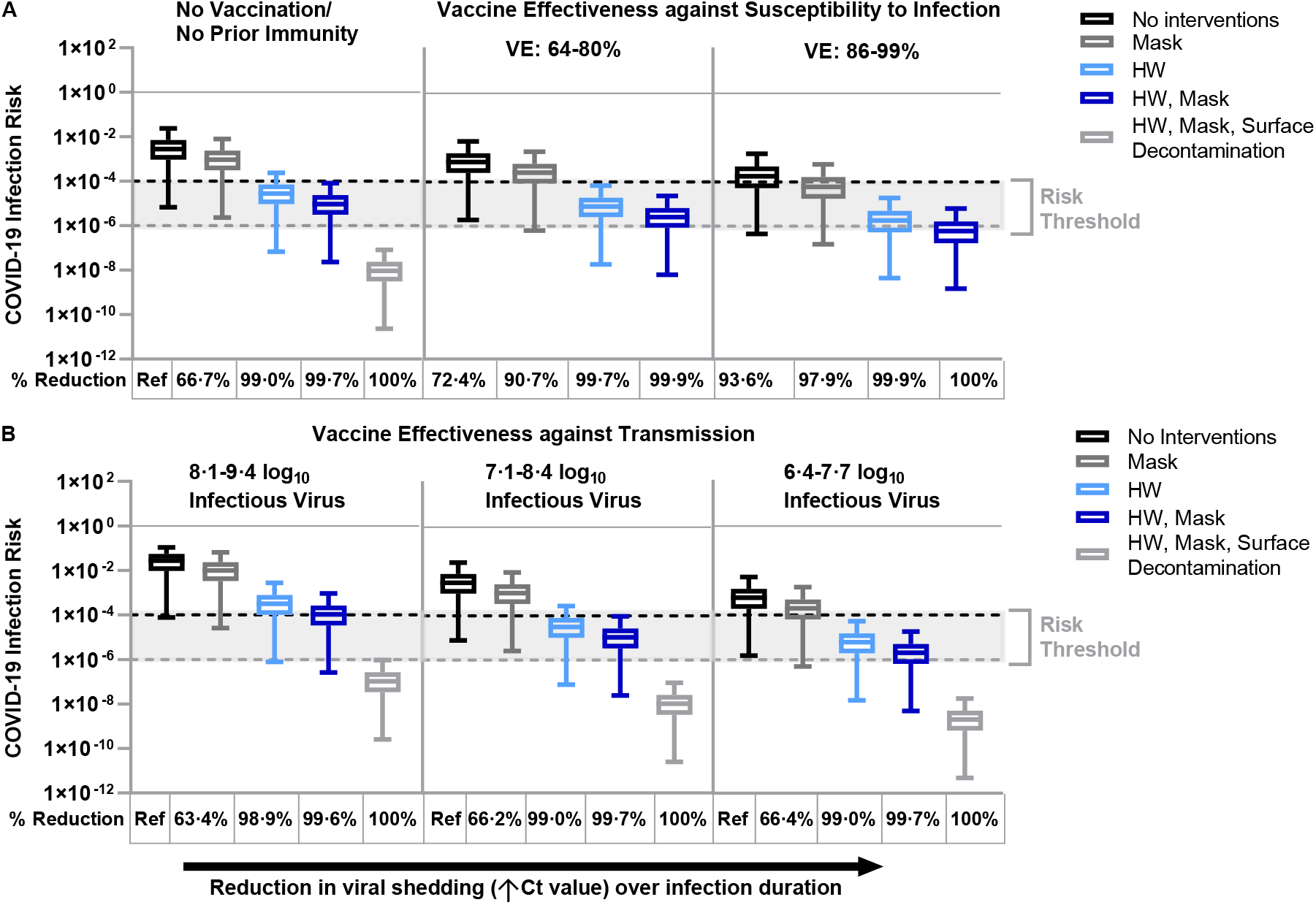
Fomite-mediated SARS-CoV-2 infection risks associated with individual and combined standard infection control measures (hourly handwashing [2 log_10_ virus removal efficiency],^58^ surgical mask use). Vaccination was incorporated into the model representing two doses of mRNA vaccine (Moderna/Pfizer) and was applied with and without the standard infection control measures. Additional decontamination of plastic packaging [3 log_10_ virus removal efficiency]^59^ was applied in combination with the standard infection control measures. Ventilation (two air changes per hour [ACH]) was applied to all simulations. An infectious to non-infectious particle ratio of 1:100^41^ was applied to all viral shedding concentrations. Reductions in SARS-CoV-2 infection risk (%) to the susceptible worker relative to no interventions are reported below each panel. Panel A represents the impact of standard infection control measures with and without vaccination on fomite-mediated SARS-CoV-2 risk. For the first vaccination scenario, we assumed only the susceptible worker was vaccinated with two doses of mRNA vaccine (Moderna/Pfizer) and vaccine effectiveness (VE) against susceptibility to infection was simulated across three vaccination states. These included: 1) no vaccination/no prior immunity; 2) lower VE ranging from 64^86^-80%^87^ representative of reduced protection (variants of concern, waning immunity, immunocompromised and elderly or at-risk populations); and 3) optimal VE ranging from 86%^88,89^-99^90^% among healthy adults 14 days or more after second mRNA dose. Panel B: the second vaccine scenario represented vaccine effectiveness against transmission, where all workers are assumed to be vaccinated with two doses of mRNA vaccines and hence the model simulated rare breakthrough infections. Vaccine effectiveness against transmission (VET) was modeled by applying the combined effect of the reduction in risk of infection to the susceptible worker and the risk of transmissibility given a rare breakthrough infection among the vaccinated workers. We used the VET estimate (88·5% [95% CI: 82·3%, 94·8%] derived from Prunas *et al*.,^62^ VET was modeled across a range of three peak infectious viral shedding concentrations representative of possible increased transmissibility and/or infectiousness of variants of concern: 1) 8·1-9·4 log_10_ viral particles; 2) 7·1-8·4 log_10_ viral particles; and 3) 6·4-7·7 log_10_ viral particles. These viral shedding levels represent 100-, 10-, and 2-times, respectively, the increased viral shedding concentration simulated in the base model analysis. Dashed lines represent 1:10,000 (black) and 1:1,000,000 (grey) infection risk thresholds, derived from WHO and U.S. EPA guidelines for drinking water quality.^60,61^

### 3.2 Impact of vaccination of the susceptible worker and infection control measures on fomite-mediated SARS-CoV-2 infection risks associated with plastic packaging under cold-chain conditions

Vaccination of the susceptible worker with two doses of mRNA vaccine, without additional infection control measures, reduced infection risk by 72·4% (reduced VE 64-80%: 7·8 × 10^−4^ risk per 1h-period, 95%CI: 1·9 × 10^−6^, 6·7 × 10^−3^), or by 93·6% (optimal VE 86-99%: 1·8 × 10^−4^ risk per 1h-period, 95%CI: 4·5 × 10^−7^, 1·8 × 10^−3^), relative to no vaccination (Figure 2A). Infection control measures in addition to vaccination of the susceptible worker (reduced VE 64-80%) further reduced risk by 90·7% (masking: 6·5 × 10^−4^ risk per 1h-period, 95%CI: 6·4 × 10^−7^, 2·3 × 10^−3^), 99·7% (hourly handwashing: 7·8 × 10^−6^ risk per 1h-period, 95%CI: 1·91 × 10^−8^, 6·8 × 10^−5^), and 99·9% (hourly handwashing and masking: 2·6 × 10^−6^ risk per 1h-period, 95%CI: 6·4 × 10^−9^, 2·3 × 10^−5^), relative to no vaccination. Optimal VE (86-99%) combined with infection control measures further enhanced the risk reduction by 93·6% (masking: 6·0 × 10^−5^ risk per 1h-period, 95%CI: 1·5 × 10^−7^, 6·2 × 10^−4^), 97·9% (hourly handwashing: 1·8 × 10^−6^ risk per 1h-period, 95%CI: 4·5 × 10^−9^, 1·9 × 10^−5^), and 99·9% (hourly handwashing and masking: 6·0 × 10^−7^ risk per 1h-period, 95%CI: 1·5 × 10^−9^, 6·2 × 10^−6^), relative to no vaccination (Figure 2A). Across all vaccination states of the susceptible worker (no vaccination/no partial immunity, reduced VE 64-80%, and optimal VE 86-99%), combined infection control measures of handwashing and masks ensured SARS-CoV-2 fomite-mediated infection risks were below 10^−4^. Notably, vaccination of the susceptible worker (VE 86-99%) combined with all standard infection control measures (handwashing, masking) resulted in risk estimates well below one in a million (10^−6^).

### 3.3 Impact of infection control measures on fomite-mediated SARS-CoV-2 infection risks with variants of concern from breakthrough cases

To account for variations in the infectiousness or transmissibility of new variants of concern (VOC), we simulated the impact of increased viral shedding concentrations resulting from rare breakthrough infections on SARS-CoV-2 fomite-mediated infection risk among a fully vaccinated workforce (two doses of Pfizer/Moderna; VET 88·5% [95% CI: 82·3%, 94·8%])^62^. Increased viral shedding (7·1-8·4 log_10_ infectious virus [10-fold increase relative to baseline shedding level]) resulted in an infection risk of 3·1 × 10^−3^ per 1h-period (95%CI: 7·8 × 10^−6^, 2·5 × 10^−2^) (Figure 2B). Implementing standard infection control measures substantially reduced risk by 99·0% for handwashing (3·2 × 10^−5^ risk per 1h-period, 95%CI: 7·8 × 10^−8^, 2·7 × 10^−4^) and by 99·7% for handwashing and masks (1·1 × 10^−5^ risk per 1h-period, 95%CI: 2·6 × 10^−8^, 9·4 × 10^−5^), relative to no infection control measures. As expected, similar trends were observed when using a 2-fold increase in infectious virus shedding, with all infection risks falling below the 10^−4^ risk threshold when implementing standard infection control measures (handwashing or handwashing and masking). In the rare event of breakthrough infections of vaccinated workers with a VOC leading to 100-fold increase in viral shedding, fomite-mediated risks remain small when continuing to use standard control measures (handwashing and masking) (*SI Appendix)*.

Additional results on the estimated SARS-CoV-2 concentration on combined plastic packaging under cold-chain conditions are located in *SI Appendix*, Figure S1.

## 4. Discussion

In this study, a stochastic QMRA model was used to quantify the relative risks of SARS-CoV-2 infection resulting from exposure to plastic fomites under cold-chain conditions in a representative frozen food packaging facility. Collectively, our modeling results indicate that risks associated with fomite-mediated transmission from plastic packaging under cold-chain conditions are extremely low, below a SARS-CoV-2 infection risk threshold of 10^−4^.^60^ Across all vaccination states of the susceptible worker (no vaccination/no prior immunity, reduced VE, optimal VE), handwashing alone or with masking resulted in risk estimates below the 10^−4^ infection risk threshold.^60^ In the case of rare breakthrough infections among fully vaccinated workers with new, potentially more infectious/transmissible variants (2- to 10-fold increased viral shedding), application of handwashing or handwashing and mask use, maintained infection risks below the 10^−4^ risk threshold. In response to recent concerns of heightened SARS-CoV-2 transmission risk resulting from exposures to contaminated plastic packaging associated with cold-chain food products,^18,19^ we found that decontaminating plastic packaging reduced risk by 3 log_10_ (9·4 × 10^−6^ vs 9·4 × 10^−9^). Given that the risk of fomite-mediated SARS-CoV-2 infection using standard infection control measures is so low, the benefit of decontaminating plastic packaging seems nominal and might be considered excessively conservative.

Exposure to plastic packaging under cold-chain conditions, even in the absence of interventions, resulted in very low fomite-mediated infection risks (under 3·0 × 10^−3^). This is consistent with Wilson *et al*., who found an infection risk associated with fomite contacts of approximately 1·0 × 10^−3^ for a single hand-to-fomite scenario with high SARS-CoV-2 bioburden and no surface disinfection.^42^ Comparable fomite-mediated risks were reported by Pitol *et al*., ^41^ and Harvey *et al*., ^26^ associated with direct tactile events in community spaces (bus stations, gas stations, playgrounds) and on high touch non-porous surfaces (crosswalk buttons, trash can handles, door handles). When implementing standard infection control measures (handwashing, mask use) and no vaccination, risk estimates ranged from 2·8 × 10^−5^ to 9·4 × 10^−6^. Aligned with our study, Pitol *et al*., also demonstrated that hand hygiene could substantially reduce the risk of SARS-CoV-2 transmission from contaminated surfaces.^41^ Our findings indicate that even for a variant for which the virus is 2- to 10-times more transmissible, fully vaccinated workers (two dose mRNA vaccine) and standard infection control measures (handwashing and masking) reduce infection risks to below the 10^−4^ risk threshold. The B.1.617.2 (Delta) SARS-CoV-2 variant should be represented within this range as recent evidence suggest it is 40-60% more transmissible than Alpha.^63,64^ This analysis demonstrates that standard infection control measures and vaccination should effectively mitigate fomite-mediated infection risks under cold-chain conditions even with highly transmissible circulating SARS-CoV-2 strains. Further, these control measures mitigate the likelihood of SARS-CoV-2 being seeded into areas in which the virus is currently not circulating.^18,19^

An important contribution of our work is quantitatively demonstrating that vaccination (susceptible worker only or full workforce), when combined with other interventions, reduced all infection risks associated with potentially contaminated frozen food packaging to below 10^−4^. Given the strong protective effect of immunity from vaccination or prior exposure, involving both reduced viral load^65^ and transmissibility,^66,67^ as more workers become vaccinated, we anticipate fomite risks will continue to decrease in occupational and community-settings alike. This holds true even when considering fomite-mediated transmission dynamics with potentially more infectious/transmissible variants, such as Delta, for which viral shedding is largely unaffected by vaccination status.^68^ Similarly, our results demonstrate that in the rare event of breakthrough infections among a fully vaccinated workforce (VET 88·5%), vaccination paired with standard infection control measures (handwashing, masking) effectively reduced risk (below 1×10^−4^). Moreover, as these analyses were conducted with a 1:100 infectious to non-infectious particle ratio, fomite-mediated transmission will be even less likely with ratios of 1:1,000–1:1,000,000, as recent studies suggest.^69-71^ Regarding reduced vaccine effectiveness (64-80%), we found fomite-mediated infection risks were reduced by 72·4% (no interventions) to 99·9% (handwashing, masking) when only the susceptible worker was vaccinated. These results are particularly promising as they encompass uncertainties in vaccine effectiveness with waning immunity ^72^ and emerging SARS-CoV-2 variants,^73,74^ heterogeneity in vaccine effectiveness and coverage across vaccine types, and variable vaccine protection among higher-risk populations (elderly, immunocompromised).^75^ It is important to note that while the global vaccine rollout continues, maintaining high compliance with mask use and handwashing will be critical given their relatively low cost, high-impact risk mitigation potential, and ease of scaling across diverse food manufacturing settings. In short, vaccination combined with masking and hand hygiene effectively minimize SARS CoV-2 transmission risk to less than 10^−4^, irrespective of additional controls and within the confines of what is currently known about variants and vaccine effectiveness.

Indeed, our study found minimal added benefit in risk reduction attributed to decontamination of plastic packaging of frozen foods. Fomite-mediated risks were found to already fall below 10^−4^ under standard infection control procedures (hourly handwashing, surgical mask use) and below 10^−6^ by adding vaccination of the susceptible worker (VE 86-99%). Risks lower than 10^−4^ already fall below U.S. EPA and WHO risk guidelines, including thresholds for drinking water (*Cryptosporidium* [9·5 × 10^−4^], *Campylobacter* [7·3 × 10^−4^], and rotavirus [2·4 × 10^−3^]) ^60^ and recreational water quality (3·2 × 10^−2^).^61^ Furthermore, surface decontamination of products meant for human consumption is not without its own risks to workers and consumers. For example, consistent occupational exposure to disinfectants is associated with adverse respiratory health outcomes, including worsening asthma control ^76^ and increased risk of chronic obstructive pulmonary disease.^77^ Also, risks to consumers of ingested disinfectants, which could enter products through damaged packaging, range in severity from irritation (nose, sinuses, skin, eyes), dizziness, and nausea to skeletal toxicity and liver damage, depending on the disinfectant type and quantity.^78,79^ Increases in the use of disinfectants since the start of the COVID-19 pandemic has resulted in a 16·4% increase in exposure calls as reported by the U.S National Poison Data System (NPDS), CDC (January–March 2019).^80^ As China is disinfecting the packaging of imported cold-chain products,^19^ future studies are warranted to investigate the adverse health effects of increased chemical exposures among workers and consumers. Furthermore, the process of testing imported frozen foods for SARS-CoV-2 and disinfecting packaging^18^ potentially introduces delays in product distribution, which could jeopardize the integrity of the products, contributing to food spoilage and waste, and could ultimately lead to shortages and instability in the global food supply chain.^81,82^ A final consideration is that increased use of disinfection products are costly, with global sales of surface disinfectant in 2020 increasing by more than 30% relative to the prior year (totaling US$4·5 billion).^7^ Notably this increased expense does not include labor costs associated with disinfecting products. In sum, additional surface decontamination of food packaging in the cold-chain could be viewed as excessive and is more likely to increase chemical risks to workers, food hazard and quality risks to consumers, and unnecessary added costs to governments and the global food industry.

Strengths of our model include a detailed exposure assessment design of frozen food packaging facilities vetted by industry and academic partners; leveraging a validated SARS-CoV-2 modeling framework;^46^ and incorporating new data as model parameters on viral persistence, fomite-to-hand transfer efficiencies, and respiratory particle definitions (aerosols and droplets). A limitation of this work is that our analysis focused on a one-hour exposure duration of the susceptible worker to contaminated plastic packaging. This duration was selected as the timing and number of products arriving in a receiving warehouse beyond one hour was unpredictable. However, when extending this to an 8-hour shift, cumulative fomite risks remained very low (handwashing and mask use: 7·5 × 10^−5^ [95%CI: 1·8 × 10^−7^-1·7 × 10^−4^]). A second limitation is that there are limited data on the concentration of the Delta variant shed by infected individuals, beyond the Ct value that estimates approximately 6 × 10^5^ copies/mL in oropharyngeal swabs.^83^ To simulate increased infectiousness/transmissibility of Delta or other VOC, we increased infectious viral shedding among the infected workers (2-, 10-, and 100-fold), which equates to 2·4 × 10^6^ to 2·4 × 10^9^ infectious virus/mL of saliva. While the Delta variant should clearly fall within or below this range of increased viral shedding, this analysis can be refined as more data become available. This would also serve to reduce the uncertainty in this and other parameters which result in a dynamic range in risk estimates and confidence intervals.

Using QMRA modeling, we find that susceptible workers (unvaccinated, no precautions) in frozen food packaging facilities are at low risk of SARS-CoV-2 fomite-mediated transmission (exposure to large droplets and small aerosol particles on plastic packaging) under cold-chain conditions. Strategies demonstrated to further reduce fomite-mediated infection risk to workers include hourly handwashing and universal mask use. Vaccination of workers should be prioritized as an effective intervention when combined with these industry standard infection control measures.^64^ When assessing the relative fomite infection risk, our work suggests that extra decontamination procedures on food packaging of cold-chain products make only negligible contributions beyond the standard infection control risk reductions. Hence, they are difficult to justify based on risk reduction. Continued use of global ^84,85^ and U.S. federal ^56,57^ SARS CoV-2 interventions (handwashing, mask use) and equitable distribution of vaccines will support the health and safety of food workers and maintain global food supply chains and consumer food security.^82^

## Data Availability

The code developed and used for this study will be made available through GitHub, with the DOI available upon publication.

## Contributors

All authors contributed to the study conceptualization and methods. JSS conducted the analysis and wrote and prepared the manuscript. All authors reviewed and revised the manuscript. JSS and DKC verified the data. All authors read and approved the final version of the manuscript. All authors had full access to study data and had final responsibility for the decision to submit for publication.

## Acknowledgments

This work was partially supported by the National Institutes of Health (NIH) T32 grant (J.S.S., grant 2T32ES012870-16), the U.S. Department of Agriculture (USDA) (J.S.L. 2018-07410; J.S.S., grant 2020-67034-31728), the National Institute General Medical Sciences (B.A.L R01 GM124280), the National Science Foundation (B.A.L. 2032084), the NIH (E.T.S., T32AI138952), and Emory University’s Infectious Disease Across Scales Training Program (E.T.S). The views expressed in this study are those of the authors and do not necessarily represent those of the NIH or USDA. The authors would like to thank Drs. Sanjay Gummalla and Lory Reveil (American Frozen Food Institute) for their valuable time and input as frozen food experts and for conducting surveys of facilities.

