## Supplemental material for "Low risk of SARS-CoV-2 transmission via fomite, even in cold-chain"

### Supplementary Information Appendix for Low risk of SARS-CoV-2 transmission via fomite, even in cold-chain.

#### Supplementary Information Appendix

##### Materials and methods:

##### Model conceptualization and vetting

Model design was informed by several frozen fruit and vegetable manufacturers and their trade association, the American Frozen Food Institute. Following initial development, the model was vetted by food industry experts: Dr. Sanjay Gummalla [Senior Vice President, Scientific and Regulatory Affairs, American Frozen Food Institute] and Dr. Lory Reveil [Director, Scientific and Regulatory Affairs, American Frozen Food Institute].

##### Data sources

The model assumed two infected workers (as unvaccinated or as rare breakthrough infections) and that the infected and susceptible workers were independent of one another. To calculate viral shedding of the infected workers, we converted PCR-based genome equivalent copies to PFU using a 1:100 conversion, as previously applied by Pitol *et al.*[1] This resulted in SARS-CoV-2 titers in saliva (range: 6·1 to 7·4 $\log_{10}$ PFU/mL),[2,3] representative of peak virus titer reported around the time of symptom onset [3,4] and the acute phase of infection, when the majority of transmission events are thought to occur.[4,5] The same distribution of shedding data was used for both infected workers. To determine the amount of virus expelled into the air by the infected workers, the total fraction of saliva volume released during coughing was calculated for each droplet (50-60µm, 60-100µm, >100µm) and aerosol (<50µm) range as described in [6] using respiratory particle counts and size distributions from empirical studies.[7]

The model simulated equal probability of cough events for each infected worker, ranging from 0-10 coughs, over the duration of product packaging (1h-period). Aerosol transport properties of the differently-sized respiratory particles informed the contamination potential of the plastic packaging. For instance, aerosols defined as <50 µm in diameter settle from the air according to their terminal settling velocity. Droplets (50-750 µm) fall rapidly due to gravitational forces and their ability to contaminate fomites as fallout or spray was determined by size and distance traveled based on modeling studies.[8] The proportion of droplets that reached the plastic cartons or plastic wrap within 0 to 3 feet distancing was derived from modeling studies by.[8]

Viral decay [9] was included at two stages in the model: 1) virus-containing aerosols or droplets in the air; and 2) the virus-contaminated hands of the susceptible worker. The model assumed no viral decay during pallet transport, holding, and unpacking while under cold chain storage conditions.[9-12] Additionally, the model assumed the susceptible worker had no additional SARS-CoV-2 exposures in the receiving warehouse (e.g., local community transmission, other infected workers in the receiving warehouse etc.,). Given heterogeneity in the volume of products processed over a contiguous workday, a 1-hour period was modeled to more precisely represent the number of products handled by workers within under cold-chain conditions. Further, the model exclusively simulated fomite-mediated transmission and did not capture potential respiratory exposures associated with re-aerosolization of virus particles from fomites. Virus transfer efficiencies from plastic fomite surfaces (individual plastic cartons, palletized cartons, plastic wrap) to hands leveraged laboratory-based studies using acrylic surfaces under low-humidity conditions and the viral surrogate MS2.[13] Sequential tactile events were modeled from the initial contact of the susceptible worker's hand to the fomite surface (one contact/individual plastic carton; up to 20 contacts on the pallet plastic wrap) followed by hand contact to facial mucous membranes (0·8 contacts/minute).[14] SARS-CoV-2 infection risks were estimated using an exponential dose-response model based on the pooled data from studies of SARS-CoV and murine hepatitis virus infection in mice by intranasal administration [15,16] with the $ID_{50}$ equal to 102 infectious particles. We applied this SARS-CoV dose-response model given the high degree of comparability to SARS-CoV-2 (e.g., genetic and amino acid homology, transmission pathways, etc.).[17]

Infection control measures were implemented to reduce contamination of the plastic packaging (i.e., mask use by the infected workers) or to disrupt the transfer of virus from the susceptible worker's hands to their mucous membranes (handwashing). Laboratory-based studies on mask filtration efficiencies [18-21] were used to inform estimates of surgical masking efficacy. Using these empirical studies, which reported mask filtration efficiencies for either source control and/or recipient protection, we calculated the mask efficacy for when the infected workers wore the mask (source) and for when the susceptible worker (recipient) wore the mask. Of note, we assumed no reduction in hand to face contacts when the susceptible worker was contacting their face. Handwashing and package surface disinfection virus removal efficiencies were representative of current CDC and EPA List N: Disinfectants for Coronavirus (COVID-19) products: handwashing (2 $log_{10}$ virus removal);[22] and plastic packaging decontamination (3 $log_{10}$ virus removal).[23] For all mitigation strategies (mask use, handwashing, and surface decontamination), we assumed that these were implemented with 100% compliance and in the specified manner. A routine ventilation rate was applied across all modeled scenarios defined as two complete room air changes per hour (ACH).

Vaccination was incorporated into the model representing two doses of mRNA vaccine (Moderna/Pfizer) and was applied with and without the standard infection control measures. For the first vaccination scenario, we assumed only the susceptible worker was vaccinated with two doses of mRNA vaccine (Moderna/Pfizer) and vaccine effectiveness (VE) against susceptibility to infection was simulated across three vaccination states. These included: 1) no vaccination/no prior immunity; 2) lower VE ranging from 64[24]-80%[25] representative of reduced protection (variants of concern, waning immunity, immunocompromised and elderly or at-risk populations); and 3) optimal VE ranging from 86%[26,27]-99[28]% among healthy adults 14 days or more after second mRNA dose. The second vaccine scenario represented vaccine effectiveness against transmission, where all workers are assumed to vaccinated with two doses of the mRNA vaccines and hence the model simulated rare breakthrough infections. Vaccine effectiveness against transmission (VET) was modeled by applying the combined effect of the reduction in risk of infection to the susceptible worker and the risk of transmissibility given a rare breakthrough infection among the vaccinated workers. We used the VET estimate (88·5% [95%CI: 82·3%, 94·8%] derived from Prunas *et al.,*[29] VET was modeled across a range of three peak infectious viral shedding concentrations representative of possible increased transmissibility and/or infectiousness of variants of concern: 1) 8·1-9·4 $log_{10}$ viral particles; 2) 7·1-8·4 $log_{10}$ viral particles; and 3) 6·4-7·7 $log_{10}$ viral particles. These viral shedding levels are 100-, 10-, and 2-times, respectively, the increased viral shedding concentration simulated in the base model analysis. For all vaccination scenarios, VE was applied directly to the model-derived risk estimates to represent reduction in infection risk. An assumption of this model is that VE would have the same impact across transmission pathways (aerosol, droplet, and fomite-mediated).

Model parameters associated with the indoor facility and respiratory transmission (aerosols and droplets) modes were described in.[30] However, air temperature was modified from 70 to 55°C to represent ambient air temperatures used during the process of freezing and packaging frozen foods. In addition, extensive sensitivity analyses including the number of simulations needed to achieve model stability; variability and uncertainty propagation throughout the model; and identifying the most influential parameters for SARS-CoV-2 infection risk, reported as Spearman's correlation coefficients, were previously described in Sobolik *et al.*[30]

**Results:**

**Impact of infection control measures on fomite-mediated SARS-CoV-2 infection risks with variants of concern from breakthrough cases**

Increased viral shedding (8·1-9·4 $log_{10}$ infectious virus [100X baseline shedding concentration]) resulted in an infection risk of 2·8 x $10^{-2}$ per 1h-period (95%CI: 7·8 x $10^{-5}$, 1·1 x $10^{-1}$) (Figure 2B). Implementing standard infection control measures reduced risk to near the $10^{-4}$ risk threshold: by 98·9% for handwashing (3·2 x $10^{-4}$ risk per 1h-period, 95%CI: 7·8 x $10^{-7}$, 2·7 x $10^{-3}$) and by 99·6% for handwashing and masks (1·1 x $10^{-4}$ risk per 1h-period, 95%CI: 2·6 x $10^{-7}$, 9·4 x $10^{-4}$), relative to no infection control measures.

**Estimated SARS-CoV-2 concentration on combined plastic packaging under cold-chain conditions**

SARS-CoV-2 concentration on combined plastic packaging in the absence of infection control measures was 11·7 infectious viruses/$m^2$ (95%CI: 2·6 x $10^{-2}$, 6·9 x $10^{1}$) (*SI Appendix*, Figure S1.A). Mask use led to a 66·7% reduction in SARS-2 concentration on fomites. The addition of plastic packaging decontamination resulted in 3·5 $log_{10}$ reduction (3·9 x $10^{-3}$ infectious viruses/$m^2$ [95%CI: 8·8 x $10^{-6}$, 2·3 x $10^{-2}$]). Handwashing by the infected workers had no impact on the fomite SARS-CoV-2 concentration as this exposure route was not considered.

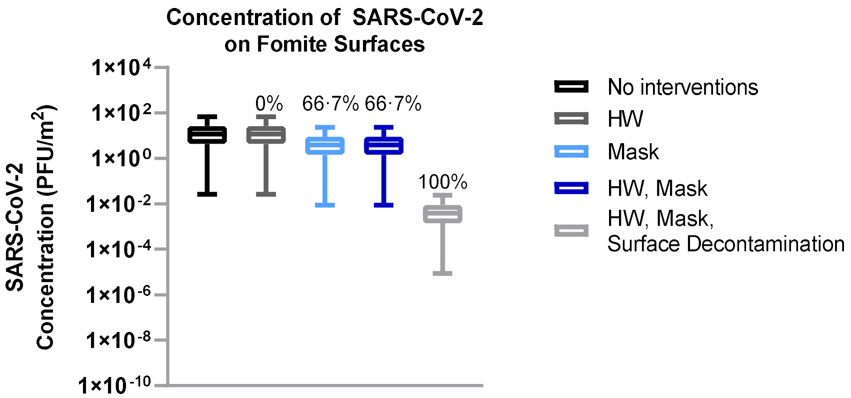


**Fig. S1. Concentration of SARS-CoV-2 on combined fomites (individual plastic cartons, plastic wrap).** SARS-CoV-2 concentrations on plastic fomite surfaces associated with standard SARS-CoV-2 infection control measures (hourly handwashing, universal surgical mask usage) under cold-chain conditions. Ventilation (two air changes per hour [ACH]) was assumed for all simulations. Percent reduction reported above each boxplot relative to no interventions.

**Table S1. Model parameter inputs and distributions.**

| Parameter | Units | Description | Distribution | Input Values | Citations |
|---|---|---|---|---|---|
| **Viral shedding** | | | | | |
| $Log_{10}(C_{virus})$ | PFU/mL | Concentration of virus in saliva<br>100X increased viral shedding<br>10X increased viral shedding<br>2X increased viral shedding | Triangular | 6·8 (6·1, 7·4)<br>8·8 (8·1, 9·4)<br>7·8 (7·1, 8·4)<br>7·1 (6·4, 7·7) | [2,3] |
| $V_{F,c}$ | mL/Cough | Fraction of volume associated with aerosols (2–45µm) | Triangular | $2{\cdot}3 \times 10^{-6}$ ($1{\cdot}4 \times 10^{-6}$, $2{\cdot}6 \times 10^{-6}$) | [7] |
| $V_{F,c}$ | mL/Cough | Fraction of volume associated with droplets (50–60µm) | Triangular | $6{\cdot}0 \times 10^{-6}$ ($3{\cdot}5 \times 10^{-6}$, $6{\cdot}7 \times 10^{-6}$) | [7] |
| $V_{F,c}$ | mL/Cough | Fraction of volume associated with droplets (60–100µm) | Triangular | $4{\cdot}9 \times 10^{-6}$ ($1{\cdot}1 \times 10^{-6}$, $8{\cdot}4 \times 10^{-6}$) | [7] |
| $V_{F,c}$ | mL/Cough | Fraction of volume associated with droplets (100–750µm) | Triangular | $6{\cdot}8 \times 10^{-3}$ ($4{\cdot}0 \times 10^{-3}$, $7{\cdot}6 \times 10^{-3}$) | [7] |
| $F_C$ | Cough/h | Number of coughs per hour | Empirical | 1/11 equal probability (0, 10) | [31,32] |
| $\lambda_{virus}$ | Hour | Viral decay of SARS-CoV-2 at 40% relative humidity, 55°F | Point value | 0·1447 | [33] |
| pp | Probability | Probability respiratory particles will remain in the air as respiratory spray between 0 and 1m distancing | Uniform | 50-60µm:<br>1m: 0·82;<br>60-100µm:<br>1m: 0·44;<br>>100µm:<br>1m: 0·04 | [8] |
| $pp_{droplets}$ | Probability | Probability respiratory particles (>100 µm) will remain in the air as respiratory spray between 0 and 1m distancing | Uniform | (0·01, 0·22) | [8] |
| $pp_{falldroplets}$ | Probability | Probability respiratory particles (>100 µm) will settle to the fomite surfaces between 0 and 1m distancing | Uniform | (0·07, 0·78) | [8] |
| **Risk mitigation interventions[1]** | | | | | |
| $S_{mask}$ | Log reduction | Source protection surgical mask efficacy | Uniform | (0·39, 0·57) | [20,34,35] |
| $RS_{mask}$ | Percent reduction | Recipient surgical mask efficacy | Uniform | (0·37, 0·998) | [20,34,35] |
| $SD_{eff}$ | Log reduction | Plastic fomite surface decontamination efficiency | Point value | $3Log_{10}$ virus | [23,36] |
| $HW_{eff}$ | Log reduction | Handwashing efficiency | Point value | $2Log_{10}$ virus | [37,38] |
| $HW_{freq}$ | Handwashing/h | Frequency of handwashing per hour | Point value | 1·0 | Expert elicitation |
| $R_{air}$ | Air changes/h | Frequency of room air changes per hour (ACH) | Point value | ACH 2 | Expert elicitation |
| $VE_{optimal}$ | Percent reduction | Vaccine effectiveness | Uniform | (0 ·86,0·99) | [26-28] |
| $VE_{reduced}$ | Percent reduction | Vaccine effectiveness | Uniform | (0·64,0·80) | [24,25] |
| VET | Percent reduction | Vaccine effectiveness against transmission | Triangular | 0·89(0·82,0·95) | [29] |
| **Fomite-mediated transmission** | | | | | |
| $SA_{carton.top}$ | $m^2$ | Surface area of top of individual plastic carton | Uniform | (0·106, 0·116) | Assumed |
| $SA_{carton}$ | $m^2$ | Surface area of a single individual plastic carton | Uniform | (0·41, 0·54) | Assumed |
| Cartons | Cartons/h | Number of individual plastic cartons processed per h | Uniform | (144, 216) | Assumed |

| | | | | | |
|---|---|---|---|---|---|
| Pallets | Pallets/h | Number of pallets processed per h | Point value | 4 | Assumed |
| $SA_{plasticwrap.side}$ | $m^2$ | Surface area of a single side of plastic wrapped pallet | Uniform | (4·2, 6·97) | Assumed |
| $SA_{plasticwrap}$ | $m^2$ | Surface area of entire plastic wrapped pallet | Uniform | (25·2, 41·8) | Assumed |
| $Fingers_{sa}$ | $m^2$ | Surface area of three finger tips touching the surface | Point value | 0·00042 | [39] |
| $H_{sa}$ | $m^2$ | Area of two hands (palms only) | Point value | 0·049 | [39] |
| $F_{decay}$ | Hour | Viral decay rate (PFU per hour) | Point value | 0·15 | [9] |
| $TE_{fh}$ | PFU | Viral transfer fraction from fomite to hand with relative humidity (40-65%); acrylic surface | Normal | 0·795 (0·212) | [13] |
| $TE_{hf}$ | PFU | Viral transfer fraction from hand to fomite surface | Point value | 0·025 | [40-43] |
| $TE_{hm}$ | PFU | Viral transfer fraction from hand to face | Normal | 0·20 (0·063) | [13] |
| freq.hs | Contacts/min | Frequency of contacts from hand to individual plastic cartons | Point value | Cartons/60 | Assumed |
| freq.hs.pw | Contacts/min | Frequency of contacts from hand to plastic wrap | Uniform | (4/60, 20/60) | Assumed |
| freq.hf | Contacts/min | Frequency of contacts from hand to face | Point value | 0 ·8 | [14] |
| $Hand_{decay}$ | Minutes | Viral decay rate on hands (PFU/min) | Uniform | (0·92, 1·47) | [41] |
| eyes.sa | $m^2$ | Surface area of mucous membranes—eyes | Uniform | ($1x10^{-5}$, $2x10^{-4}$) | [44] |
| nose.sa | $m^2$ | Surface area of mucous membranes—nose | Uniform | ($1x10^{-5}$, $1x10^{-3}$) | [44] |
| mouth.sa | $m^2$ | Surface area of mucous membranes—mouth | Uniform | ($1x10^{-4}$, $4.1x10^{-3}$) | [44] |
| **SARS-CoV-2 dose and risk characterization** | | | | | |
| $Ratio_{infectious}$ | No units | Infectious to non-infectious ratio | Point value | 1:100 | [45] |
| $k_{risk}$ | $PFU^{-1}$ | Dose-response parameter | Point value | 0·00680 | [1] |

[1]All interventions were assumed to be implemented with 100% compliance.
